# Development and Validation of VC-MAES and VC-SEPS: Deep Learning-Based Early Warning Systems for Hospitalized Patients

**DOI:** 10.1101/2025.08.20.25334022

**Authors:** Yeji Kim, Sangchul Hahn, Kwang Joon Kim, Eunho Yang, Ji-hyun Kim, Suji Lee, Chang Hoon Han, Joo-Yun Won, Byung Eun Ahn, Yechan Mun, Kyung Soo Chung, Taeyong Sim

## Abstract

The timely detection of ward deterioration—including unplanned intensive care unit (ICU) transfer, cardiac arrest, death, and sepsis—remains an unmet need. Although rule-based early warning scores and newer machine-learning models have been introduced, their clinical adoption is limited owing to challenges such as low predictive performance, excessive false alarms, and lack of generalizability.

This study aimed to develop two deep-learning models for patients in general wards using a bidirectional long short-term memory neural network architecture: (i) the VitalCare-Major Adverse Event Score (VC-MAES), which predicts clinical deterioration events (CDEs)— unplanned ICU transfer, cardiac arrest, or in-hospital death—within 6 h and (ii) the VitalCare-SEPsis Score (VC-SEPS), which predicts sepsis onset within 4 h. Additionally, we sought to externally validate the performance of the models in an independent cohort.

This study was conducted in two sequential phases. First, the VC-MAES and VC-SEPS models were developed using a large retrospective cohort from Yonsei Severance Hospital, Seoul, Republic of Korea. Second, external validation was performed in a single-center cohort at National Health Insurance Service Ilsan Hospital (NHIS Ilsan Hospital), Ilsan, Republic of Korea. Both algorithms incorporated patient age, vital signs, laboratory results, and the Glasgow Coma Scale scores. VC-MAES performance was compared with the Modified Early Warning Score (MEWS) and National Early Warning Score (NEWS), whereas VC-SEPS performance was compared with the Sequential Organ Failure Assessment (SOFA), quick SOFA (qSOFA), and NEWS, using the area under the receiver operating characteristic curve (AUROC) as the primary performance metric.

The derivation cohort comprised 357,009 adult general-ward admissions at Yonsei Severance Hospital (2013–2017), and the external validation cohort included 22,073 admissions at NHIS Ilsan Hospital (2017). In the external validation cohort, the VC-MAES predicted CDEs within 6 h with an AUROC of 0.918 (95% confidence interval [CI], 0.909–0.927), outperforming the MEWS (0.834; 95% CI, 0.820–0.849) and NEWS (0.883; 95% CI, 0.869– 0.896). VC-SEPS predicted sepsis onset within 4 h, with an AUROC of 0.941 (95% CI, 0.934–0.947), surpassing the SOFA (0.559; 95% CI, 0.546–0.571), qSOFA (0.687; 95% CI, 0.671–0.704), and NEWS (0.767; 95% CI, 0.748–0.785). Both models maintained AUROC values above 0.86 across all age and sex categories.

The VC-MAES and VC-SEPS outperformed conventional early warning scores in predicting clinical deterioration events and sepsis. These models can enable earlier and more precise interventions, enhancing patient care and optimizing resource use, ultimately leading to better patient outcomes without overwhelming healthcare providers.

## Introduction

Clinical deterioration, occurring in up to 5% of hospitalized patients, is associated with increased morbidity, mortality, prolonged hospital stays, and higher healthcare costs (1–4). Among life-threatening conditions encountered in general wards (GWs), sepsis is one of the most fatal (5, 6). Therefore, timely recognition and intervention are essential to improving patient outcomes (7–11). However, early signs of clinical deterioration are often subtle and heterogeneous, making prompt recognition difficult (12, 13). Unlike intensive care units (ICUs) with continuous monitoring, GWs rely on intermittent measurements and complex communication workflows, leading to delayed critical interventions (14, 15). These cumulative constraints introduce critical time lags in recognizing and managing deterioration, directly contributing to poor outcomes (16–19).

To address these challenges, many hospitals have implemented Early Warning Scores (EWS) and Rapid Response Systems, with mixed success obtained with these interventions (20–23). EWS such as the Modified Early Warning Score (MEWS) and National Early Warning Score (NEWS) allow the prompt identification of clinical deterioration using predefined thresholds of vital signs. More recently, machine learning (ML) models have been developed to improve predictive performance by capturing complex, non-linear relationships in large-scale clinical data (24, 25).

Despite promising results, the clinical adoption of ML-based prediction tools remains limited (26). Common barriers include the lack of external validation, high false-alarm rates contributing to alarm fatigue, and insufficient subgroup analyses for fairness and bias assessment (27–30). These factors can undermine clinician trust and reduce real-time clinical utility.

To overcome these limitations, we developed two deep learning (DL)-based prediction models for use in GWs: the VitalCare Major Adverse Event Score (VC-MAES), which predicts unplanned ICU transfer, cardiac arrest, or death within 6 h, and the VitalCare-SEPsis Score (VC-SEPS), which predicts the onset of sepsis within 4 h. These models are based on recurrent neural networks, a DL architecture particularly suitable for physiological applications that process sequential data. This architecture allows the models to effectively analyze longitudinal time-series data, revealing context-dependent patterns overlooked by rule-based scores.

The objectives of this study were twofold: (1) to develop multivariable prediction models for the early identification of clinical deterioration events (CDEs) and sepsis in GW patients and (2) to validate the performance of the prediction models against a prespecified area under the receiver operating characteristic curve (AUROC) target of 0.80 through retrospective validation in an external single-center cohort. We compared the VC-MAES and VC-SEPS with established EWS and clinical scoring systems to assess whether our models enhance predictive discrimination. Furthermore, subgroup analyses across key demographic features, including age and sex, were conducted to assess performance consistency and mitigate potential bias.

## Materials and Methods

### Derivation Cohort

This study comprised two sequential phases. Phase 1 involved the development of two DL-based early-warning models, the VC-MAES and VC-SEPS, utilizing a large retrospective derivation cohort from Yonsei Severance Hospital, a tertiary care hospital in Seoul, the Republic of Korea. Adult patients aged ≥19 years with GW admissions between January 2013 and December 2017 were included. Patients were excluded if they met any of following criteria: (1) missing at least one recorded measurement for systolic blood pressure (SBP), diastolic blood pressure (DBP), heart rate (HR), respiratory rate (RR), and body temperature and (2) the occurrence of a CDE during an ICU stay. For both models, hourly time-series data of vital signs and contemporaneous laboratory results were harmonized onto a standardized hourly grid. These data were retrospectively linked with outcome labels specific to each model: CDEs for the VC-MAES and sepsis for the VC-SEPS. Detailed definitions and criteria for outcome labeling are provided in the Outcomes and Reference Standards subsection.

### Validation Cohort

Phase 2 of the study involved a single-center external validation at the National Health Insurance Service Ilsan Hospital (NHIS Ilsan Hospital), a secondary care hospital in Ilsan, Republic of Korea. A retrospective cohort comprising all adult general ward admissions between January 1, 2017 and December 21, 2017 was assembled, and this dataset was used to externally validate the two deep learning–based models developed in Phase 1.

### Outcomes and Reference Standards

For the VC-MAES model, the composite outcome was defined as any unplanned ICU transfer—specifically, a direct transfer from a general ward to the ICU (34)—in-hospital cardiac arrest confirmed by chest compressions or defibrillation (35), or in-hospital death occurring during the index admission; these outcomes are commonly used to define clinical deterioration in hospitalized patients (36, 37). For unplanned ICU transfers, patients with a history of surgery were included only if the interval between the end of the surgery and ICU admission exceeded 24 h, since transfers within 24 h are more likely to represent planned postoperative ICU stays rather than true clinical deterioration.

For the VC-SEPS model, sepsis was defined based on the Sepsis-3 consensus as “life-threatening organ dysfunction caused by a dysregulated host response to infection” (5). Operationally, however, we applied the CDC’s Adult Sepsis Event (ASE) toolkit, a surveillance-oriented definition derived from Sepsis-3 but simplified and optimized for reliable use with routinely collected electronic health record (EHR) data (38–40).

Specifically, ASE defines sepsis as the concurrent presence of a suspected infection, indicated by the collection of blood cultures, and antibiotic administration for at least four consecutive qualifying antimicrobial days (QADs). Instead of relying on an increased Sequential Organ Failure Assessment (SOFA) score, ASE employs a simplified definition of acute organ dysfunction, characterized by the initiation of vasopressor therapy or mechanical ventilation, elevated lactate levels, or alterations in creatinine, total bilirubin, or platelet counts compared to clearly established baseline values (39). The simplified version intentionally excludes variables frequently measured, documented, or recorded inconsistently within EHRs, such as mental status assessments, vasopressor dosages, urine output, blood gas data, and FiO₂ values recorded concurrently with blood samples. By omitting these less consistently documented elements, the ASE definition achieves greater objectivity and enhanced applicability across diverse hospital settings and varying EHR systems (41, 42). In addition, the requirement of at least 4 QADs minimizes false-positive cases arising from brief courses of empiric antibiotics that are subsequently discontinued when infection is no longer suspected (39, 43–45).

Because both event types (CDEs for VC-MAES and ASE-defined sepsis) are captured via structured, time-stamped fields within the EHR, outcome determination was objective and required no adjudicator masking.

### Predictors

Both algorithms primarily utilize five core vital signs—SBP, DBP, HR, RR, and body temperature—as well as patient age. When available, additional clinical parameters, such as peripheral oxygen saturation, the Glasgow Coma Scale (GCS), total bilirubin, lactate, creatinine, platelet count, arterial pH, sodium, potassium, hematocrit, white blood cell count, bicarbonate, and C-reactive protein, are incorporated into the models to enhance comprehensive risk prediction. Notably, in cases where the GCS was not directly recorded, documented mentions of the consciousness level of a patient in their medical records were converted to corresponding GCS scores. All clinical inputs were aligned onto a standardized hourly grid, enabling consistent temporal data representation. The DL-based network subsequently computed and produced a continuous risk score of 0–100, reflecting the risk of CDEs or sepsis.

### Missing Data

Intermittent gaps in clinical data were managed using the last observation carried forward method, which imputes missing values using the most recent prior measurement. This method is commonly employed in clinical prediction models using longitudinal EHR data, as it preserves temporal continuity and clinical plausibility by assuming relative stability between measurements (46, 47). When an entire variable channel was absent—for example, lactate levels were never drawn—the midpoint of the clinically accepted normal reference range was imputed at the earliest timestamp and subsequently propagated forward. This approach reflects the clinical assumption that missing measurements often indicate a normal physiological state, as clinicians typically measure a variable only when abnormality is suspected (48). This combined strategy was supported by internal benchmarking, demonstrating preservation of predictive discrimination, alignment with clinical reasoning, and maintenance of physiological plausibility. Reference midpoints or normal values used for imputing missing variables are provided in **S1 Table**.

### Ethics

The Institutional Review Board (IRB) of Yonsei Severance Hospital (Approval No. 4-2017-0939) waived ethical approval for the derivation cohort of this work because only the de-identified electronic health record data were used. The IRB of NHIS Ilsan Hospital (Approval No. NHIMC 2018-06-004-001) gave ethical approval for the external validation cohort of this work and waived the requirement for informed consent. All data collection adhered to the principles of the Declaration of Helsinki.

### Model Development

The development of the VC-MAES and VC-SEPS predictive models employed an advanced DL-based approach designed specifically for binary classification tasks involving clinical time-series data. Both models were based on a bidirectional long short-term memory (biLSTM) neural network architecture, optimized to capture temporal dependencies inherent in sequential clinical data.

The input data for both models were categorized into dynamic and static features. Dynamic features comprised hourly collected clinical measurements, and static features included demographic and clinical baseline characteristics.

The biLSTM component processed dynamic inputs, using its ability to capture temporal relationships by analyzing clinical measurements both forward and backward through time. Static features were handled separately by fully connected neural network layers (deep neural networks, DNNs).

Outputs from both biLSTM and fully connected layers were integrated into a unified latent representation. Afterwards, this merged representation was processed through additional fully connected classification layers, resulting in a binary prediction output indicating the risk of CDE (VC-MAES) or sepsis (VC-SEPS).

Model training utilized retrospective EHRs, employing a standardized approach for missing data to handle intermittent gaps and entirely absent variable channels. Final model predictions were expressed as continuous risk scores of 0–100, with predefined threshold values to guide clinical interventions. The overall model architecture is illustrated in Fig 1.

**Fig 1.**
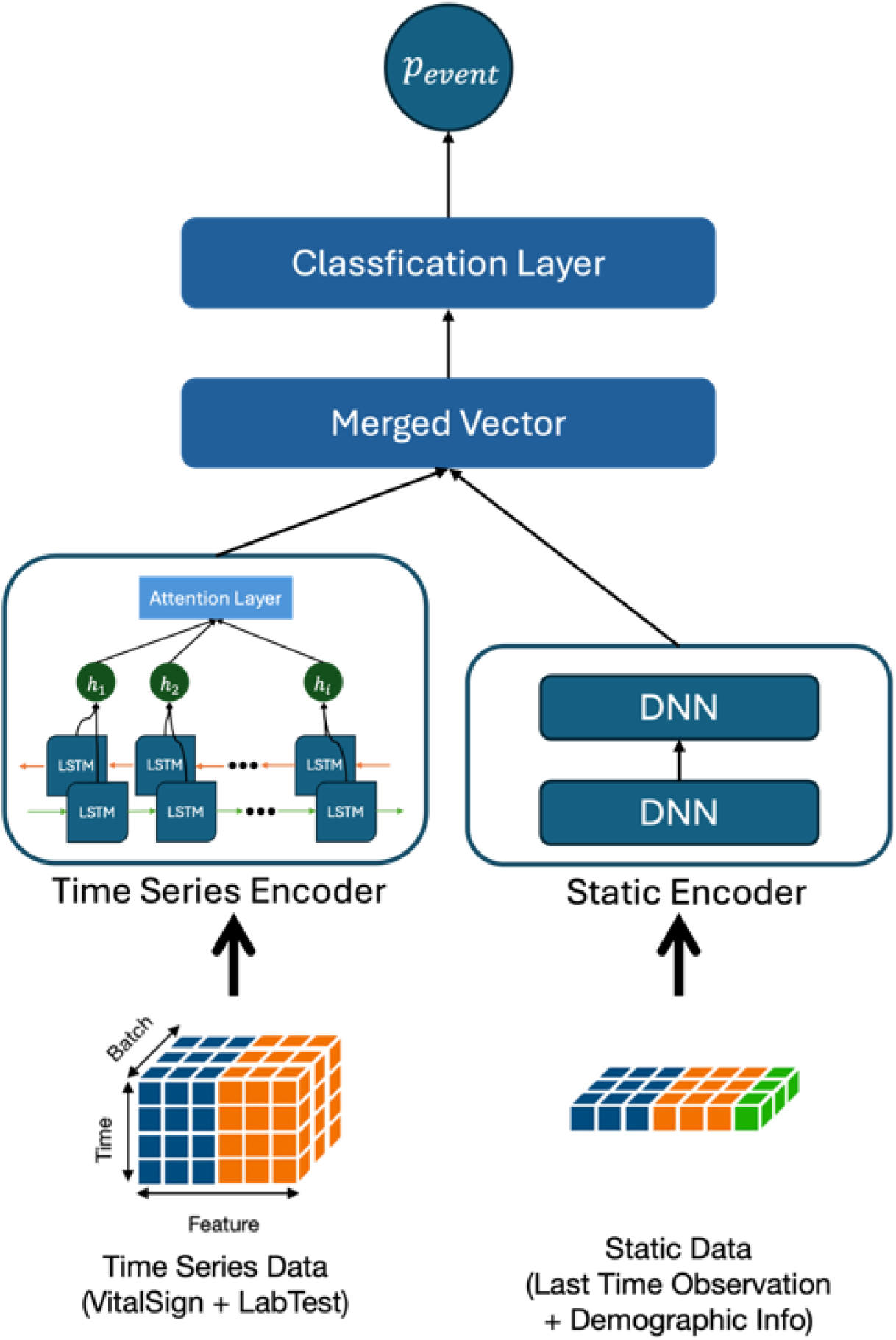
Overall model architecture of the VC-MAES and VC-SEPS. The models process dynamic features using a biLSTM (bidirectional long short-term memory) followed by an attention layer, and static features using DNNs (deep neural networks). The outputs are integrated and passed through classification layers to predict the risk of clinical deterioration (VC-MAES) or sepsis (VC-SEPS).

### Subgroup Analysis

To assess model performance across key demographic subpopulations, we conducted a stratified subgroup analysis by sex (male vs. female) and age group. For the age-based analysis, patients were grouped into predefined 10-year strata from 19 to 90 years (19–29, 30–39, 40–49, 50–59, 60–69, 70–79, and ≥80 years), and performance metrics were assessed within each age group. For the sex-based analysis, model performance was evaluated separately for male and female patients. This approach was used to examine the fairness and consistency of model performance across clinically relevant demographic subgroups.

### Performance Evaluation and Statistical Analysis

The primary performance measure used for evaluating the VC-MAES and VC-SEPS algorithms was the AUROC. The AUROC effectively quantifies the ability of a model to discriminate between event-positive (CDEs or sepsis) and event-negative cases (49). An AUROC value of 0.8 or higher was predefined as the minimum acceptable standard for clinical utility (31). For statistical comparison of AUROC values, the DeLong’s test was used; the VC-MAES was compared with the MEWS and NEWS, and the VC-SEPS was compared with SOFA, quick SOFA (qSOFA), and NEWS.

The area under the precision-recall curve (AUPRC) was assessed as a key complementary metric for evaluating prediction models, especially in the context of rare events (50, 51).

Additionally, the predictive scores of the models were translated into binary classifications using predefined threshold values, enabling practical use in clinical decision-making.

Sensitivity, specificity, positive predictive value (PPV), and negative predictive value (NPV) were calculated to provide comprehensive performance metrics, supporting robust assessment of clinical applicability.

Owing to their extensive use worldwide for predicting cardiac arrest and clinical deterioration (52, 53), the NEWS and MEWS served as comparative measures for VC-MAES in this study. Similarly, based on their utility as sepsis prognosis markers validated in multiple studies, qSOFA, SOFA, and NEWS were used as comparative measures for the VC-SEPS (54).

All statistical analyses, including the calculation of AUROC and confidence intervals, were performed using Python software (version 3.6 or higher). Statistical significance was defined using a two-sided alpha level of 0.05.

## Results

### Participants and Baseline Characteristics

The overall study flowchart is shown in Fig 2. Both the VC-MAES and VC-SEPS models were developed and validated using the same derivation and validation cohorts. In the derivation cohort from Yonsei Severance Hospital, 370,096 hospitalizations were initially screened; no patients were younger than 19 years, and 13,087 encounters were excluded according to the predefined exclusion criteria, resulting in 357,009 hospitalizations involving 203,977 patients. From this cohort, 283,040 and 73,969 encounters were assigned to the training and internal test sets for VC-MAES, respectively, while 284,340 and 72,669 encounters were assigned to the training and internal test sets for VC-SEPS. In the validation cohort from NHIS Ilsan Hospital, 31,913 hospitalizations were screened; 4,264 admissions involved patients younger than 19 years, and 5,576 were excluded according to the exclusion criteria, yielding 22,073 hospitalizations involving 17,188 patients.

**Fig 2.**
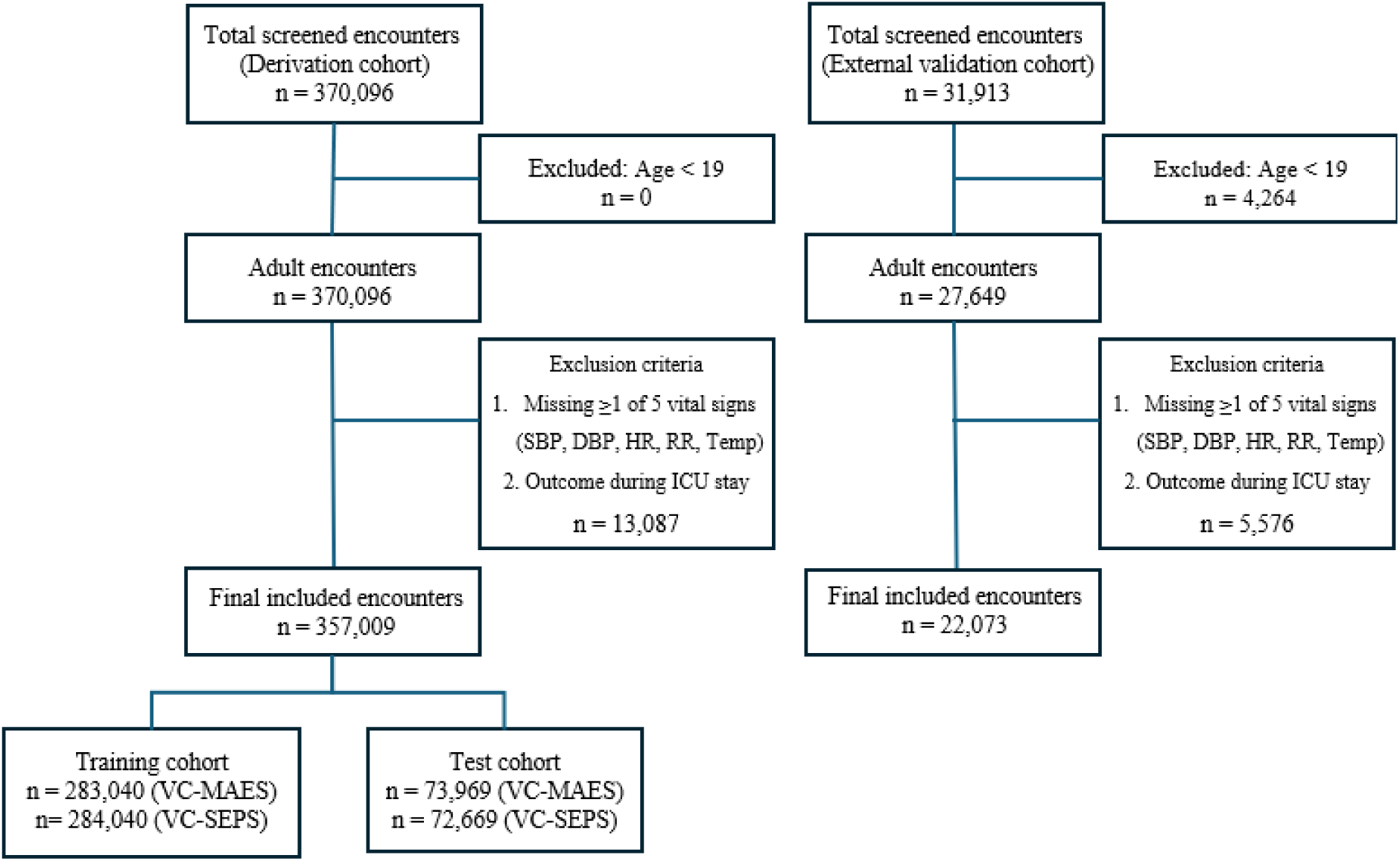
Study Flowchart. Flowchart depicting patient selection, exclusions, and final sample sizes for the derivation and external validation cohorts. ICU = intensive care unit.

Table 1 presents patient demographics, comorbidities, and the median values of model input features. The validation cohort had a consistently higher burden across all comorbidities included in the Charlson Comorbidity Index. In addition, the validation cohort showed a higher prevalence of sepsis (3.7% vs. 2.2%) and in-hospital mortality (2.9% vs. 1.1%) compared to the derivation cohort. The incidence of in-hospital cardiac arrest remained identical at 0.2% in both cohorts. In contrast, the overall rate of CDEs was lower in the validation cohort (4.6% vs. 6.0%), largely driven by a markedly lower rate of ward-to-ICU transfer (1.4% vs. 4.7%). This is particularly significant because ward-to-ICU transfers represent the majority of CDE cases.

**Table 1.** Baseline demographic characteristics, comorbidities, vital signs, laboratory test values, length of stay, of derivation and validation cohort.

| Group | Derivation cohort<br>(N=357,009) | External validation cohort<br>(N=22,073) |
| --- | --- | --- |
| N. of patients |  |  |
| N. of hospitalizations | 357,009 | 22,073 |
| N. of subjects | 203,977 | 17,188 |
| Mean age (mean ± SD), y | 56.1 ± 16.3 | 60.8 ± 17.9 |
| Sex (%) |  |  |
| - Male | 176,738 (49.5%) | 10,387 (47.1%) |
| - Female | 180,271 (50.5%) | 11,686 (52.9%) |
| **Comorbidities (%)** |  |  |
| Myocardial infarction | 438 (0.1 %) | 85 (0.4 %) |
| Chronic heart failure | 1,634 (0.5 %) | 466 (2.1 %) |
| Peripheral vascular disease | 1,811 (0.5 %) | 323 (1.5 %) |
| Cerebrovascular disease | 4,130 (1.2 %) | 1,170 (5.3 %) |
| Dementia | 975 (0.3 %) | 293 (1.3 %) |
| Chronic pulmonary disease | 2,324 (0.7 %) | 684 (3.1 %) |
| Rheumatic disease | 521 (0.1 %) | 155 (0.7 %) |
| Peptic ulcer disease | 1,597 (0.4 %) | 237 (1.1 %) |
| Mild liver disease | 2,635 (0.7 %) | 852 (3.9 %) |
| Diabetes without complication | 6,524 (1.8 %) | 1,539 (7.0 %) |
| Diabetes with complication | 908 (0.3 %) | 514 (2.3 %) |
| Paraplegia and hemiplegia | 989 (0.3 %) | 469 (2.1 %) |
| Renal disease | 4,609 (1.3 %) | 1,660 (7.5 %) |
| Any malignancy | 30,456 (8.5 %) | 4,336 (19.6 %) |
| Moderate or severe liver disease | 623 (0.2 %) | 106 (0.5 %) |
| Metastatic carcinoma | 8,988 (2.5 %) | 675 (3.1 %) |
| AIDS/HIV | 50 (0.0 %) | 12 (0.1 %) |
| **Primary variables for VC-MAES and VC-SEPS (mean ± SD)** |  |  |
| SBP (mmHg) | 126.76±18.78 | 129.31±20.65 |
| DBP (mmHg) | 78.33±12.08 | 76.44±13.47 |
| HR (beat/min) | 79.77±15.21 | 81.87±15.73 |
| RR (breathes/min) | 19.21±1.83 | 19.25±2.79 |
| Body temperature (°C) | 36.75±0.48 | 36.92±0.55 |
| **Additional variables for VC-MAES and VC-SEPS (mean ± SD)** |  |  |
| Oxygen saturation (%) | 96.98±4.81 | 95.72±6.60 |
| Glasgow Coma Scale | 14.95±0.42 | 13.23±1.72 |
| Total bilirubin (mg/dL) | 1.32±2.60 | 1.21±2.10 |
| Lactate (mmol/L) | 1.44±1.25 | 2.96±2.77 |
| pH | 7.42±0.05 | 7.43±0.07 |
| Sodium (mEq/L) | 139.13±3.97 | 137.14±4.49 |
| Potassium (mEq/L) | 4.25±0.52 | 4.11±0.56 |
| Creatinine (mg/dL) | 1.20±1.59 | 1.26±1.74 |
| Hematocrit (%) | 35.87±5.89 | 36.49±6.29 |
| WBC (10<sup>9</sup>/L) | 7.99±4.67 | 8.87±4.75 |
| Bicarbonate (mEq/L) | 21.43±3.45 | 23.11±4.81 |
| Platelet (10<sup>9</sup>/L) | 225.98±96.49 | 234.97±90.62 |
| CRP (mg/dL) | 38.56±59.54 | 48.81±64.98 |
| **Length of stays [95% CI]** |  |  |
| Median [IQR], days | 4 [2, 8] | 5 [3, 11] |
| **No. of outcomes (%)** |  |  |
| Outcome of VC-SEPS |  |  |
| Sepsis | 7,865 (2.2%) | 820 (3.7%) |
| Outcomes of VC-MAES | 21,428 (6%) | 1,020 (4.6%) |
| Ward to ICU transfer | 16,751 (4.7%) | 315 (1.4%) |
| Mortality | 4,007 (1.1%) | 651 (2.9%) |
| IHCA | 670 (0.2%) | 54 (0.2%) |

CI, Confidence Interval; AIDS/HIV, Acquired Immunodeficiency Syndrome/Human Immunodeficiency Virus; SBP, Systolic Blood Pressure; DBP, Diastolic Blood Pressure; HR, Heart rate; RR, Respiratory Rate; CRP, C-reactive protein; WBC, White Blood Cell; VC-SEPS, VitalCare-SEPs Score; VC-MAES, VitalCare-Major Adverse Event Score; ICU, Intensive care unit; IHCA, In-Hospital Cardiac Arrest.

### Missing Rates

There was a substantial variation in missing data patterns between the derivation and validation cohorts (S2 Table). The GCS showed a substantially higher missing rate in the validation cohort (98.1%) compared to the derivation cohort (12.6%). This significant discrepancy in missing rates suggests that routine documentation and assessment of consciousness level were more frequently performed in the derivation cohort. Conversely, SpO₂ exhibited a lower missing rate in the validation cohort (45.7%) than in the derivation cohort (66.7%). Similarly, for most laboratory features—excluding pH, lactate, bicarbonate— the validation cohort showed lower missing rates than did the derivation cohort. Bicarbonate was more routinely measured in the derivation cohort (78.9%) than in the validation cohort (88.9%), as reflected by its lower missing rate in the derivation cohort. Both pH and lactate levels, typically obtained via arterial or venous blood gas analysis, exhibited high missingness across both sites, reflecting limited routine use in GW settings.

### Predictive Performance

In the derivation and external validation cohorts, the VC-MAES showed significantly higher AUROCs than did the MEWS and NEWS (all p < 0.001), and the VC-SEPS showed significantly higher AUROCs than did the qSOFA, SOFA, and NEWS (all p < 0.001).

Detailed AUROC and AUPRC values are presented in Table 2, and the corresponding AUROC and AUPRC curves are shown in Fig 3. Both VC-MAES and VC-SEPS exceeded the predefined performance threshold (AUROC ≥ 0.80) and consistently outperformed conventional EWSs across cohorts. Detailed performance metrics at key cut-off values in the external validation cohort for VC-MAES and VC-SEPS are shown in S3 Table.

**Fig 3.**
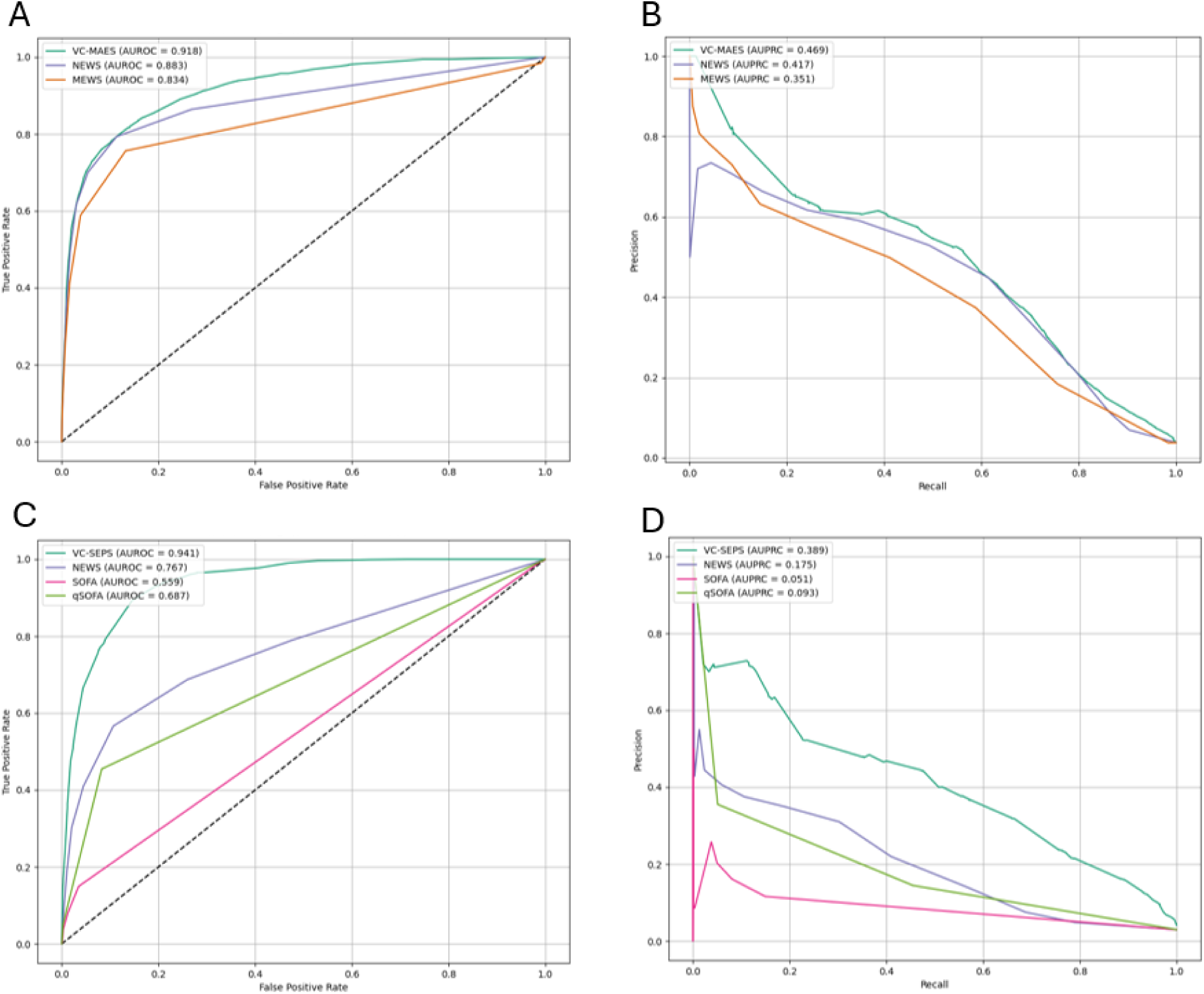
ROC and PRC curves in the external validation cohort: (A) ROC curves for clinical deterioration prediction within a 6-h window: VC-MAES vs. MEWS and NEWS (B) PRC curves for clinical deterioration prediction within a 6-h window: VC-MAES vs. MEWS and NEWS (C) ROC curves for sepsis prediction within a 4-h window: VC-SEPS vs. SOFA, qSOFA, and NEWS (D) PRC curves for sepsis prediction within a 4-h window: VC-SEPS vs. SOFA, qSOFA, and NEWS ROC, Receiver Operating Characteristic; PRC, Precision-Recall Curve; VC-MAES, Vital Care-Major Adverse Event Score; MEWS, Modified Early Warning Score; NEWS, National Early Warning Score; VC-SEPS, VitalCare-SEPsis Score; SOFA, Sequential Organ Failure Assessment; qSOFA, quick Sequential Organ Failure Assessment

**Table 2.**
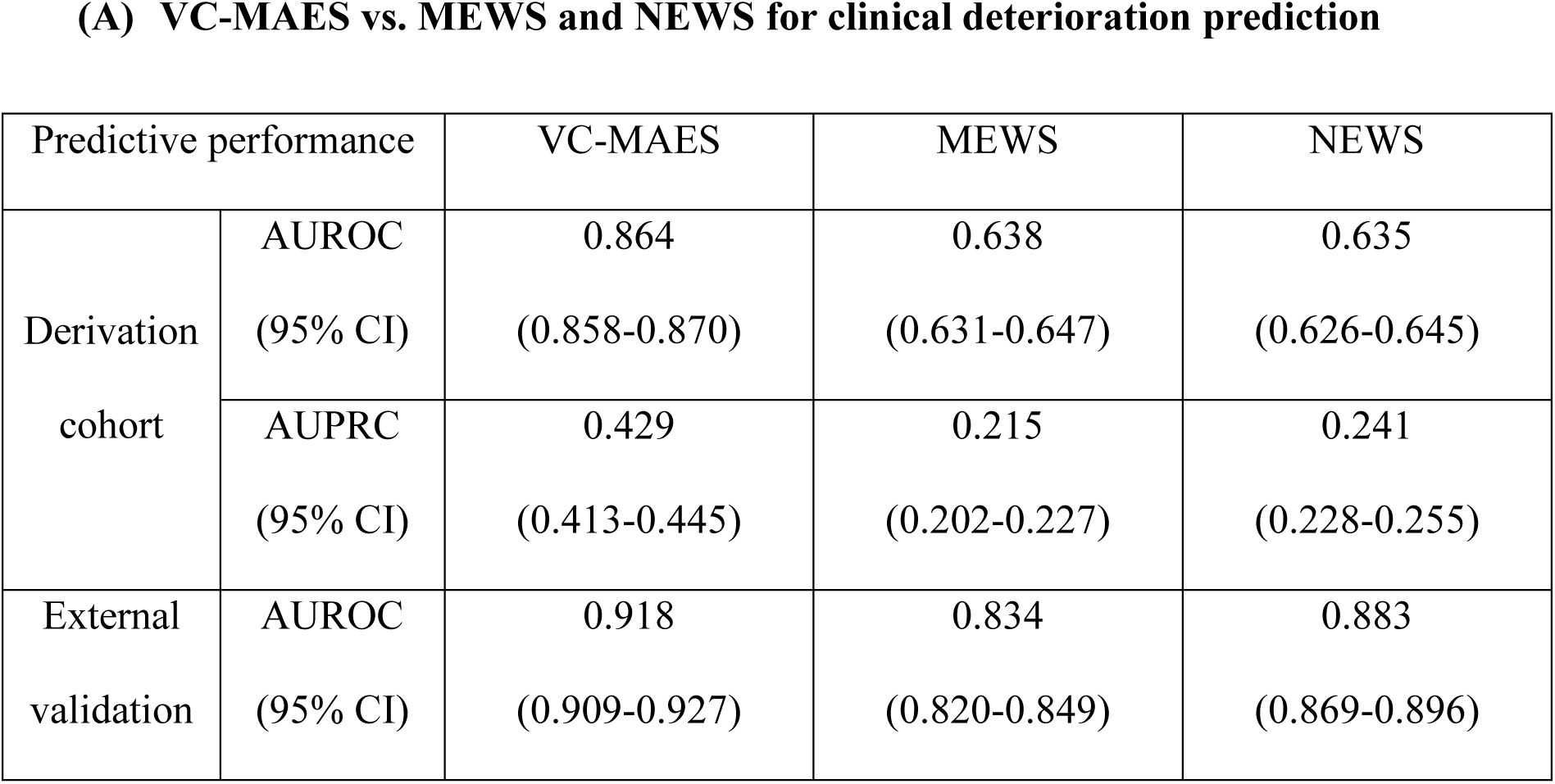
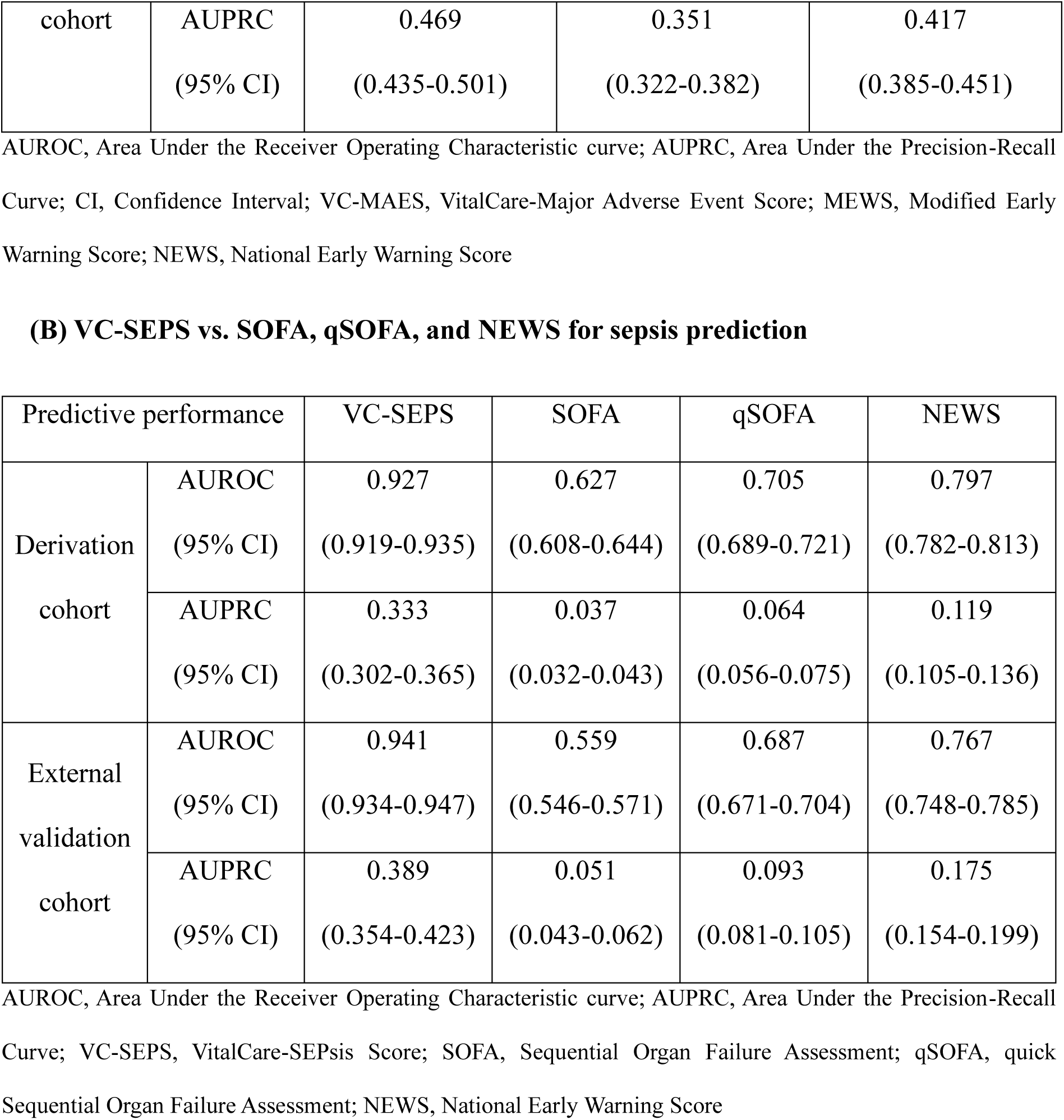
AUROC and AUPRC of risk scores for predicting clinical deterioration within 6 h and sepsis within 4 h in the derivation and external validation cohorts.

(A) VC-MAES vs. MEWS and NEWS for clinical deterioration prediction
| Predictive performance |  | VC-MAES | MEWS | NEWS |
| --- | --- | --- | --- | --- |
| Derivation cohort | AUROC | 0.864 | 0.638 | 0.635 |
|  | (95% CI) | (0.858-0.870) | (0.631-0.647) | (0.626-0.645) |
|  | AUPRC | 0.429 | 0.215 | 0.241 |
|  | (95% CI) | (0.413-0.445) | (0.202-0.227) | (0.228-0.255) |
| External validation | AUROC | 0.918 | 0.834 | 0.883 |
|  | (95% CI) | (0.909-0.927) | (0.820-0.849) | (0.869-0.896) |
| cohort | AUPRC | 0.469 | 0.351 | 0.417 |
|  | (95% CI) | (0.435-0.501) | (0.322-0.382) | (0.385-0.451) |

AUROC, Area Under the Receiver Operating Characteristic curve; AUPRC, Area Under the Precision-Recall Curve; CI, Confidence Interval; VC-MAES, VitalCare-Major Adverse Event Score; MEWS, Modified Early Warning Score; NEWS, National Early Warning Score

**(B) VC-SEPS vs. SOFA, qSOFA, and NEWS for sepsis prediction**
| Predictive performance |  | VC-SEPS | SOFA | qSOFA | NEWS |
| --- | --- | --- | --- | --- | --- |
| Derivation cohort | AUROC | 0.927 | 0.627 | 0.705 | 0.797 |
|  | (95% CI) | (0.919-0.935) | (0.608-0.644) | (0.689-0.721) | (0.782-0.813) |
|  | AUPRC | 0.333 | 0.037 | 0.064 | 0.119 |
|  | (95% CI) | (0.302-0.365) | (0.032-0.043) | (0.056-0.075) | (0.105-0.136) |
| External validation cohort | AUROC | 0.941 | 0.559 | 0.687 | 0.767 |
|  | (95% CI) | (0.934-0.947) | (0.546-0.571) | (0.671-0.704) | (0.748-0.785) |
|  | AUPRC | 0.389 | 0.051 | 0.093 | 0.175 |
|  | (95% CI) | (0.354-0.423) | (0.043-0.062) | (0.081-0.105) | (0.154-0.199) |

AUROC, Area Under the Receiver Operating Characteristic curve; AUPRC, Area Under the Precision-Recall Curve; VC-SEPS, VitalCare-SEPSis Score; SOFA, Sequential Organ Failure Assessment; qSOFA, quick Sequential Organ Failure Assessment; NEWS, National Early Warning Score

### Subgroup Analysis

Table 4 presents the age- and sex-stratified AUROC comparisons for the VC-MAES and VC-SEPS in the external validation cohort. For clinical deterioration prediction (Table 4A), the VC-MAES (AUROC range, 0.861–0.998) consistently outperformed the MEWS (0.783–0.958) and NEWS (0.807–0.990) in all age and sex categories. The highest AUROC was observed in patients aged 30–39 years (0.998; 95% CI, 0.994–1.000) and lowest in those aged 40–49 years (0.861; 95% CI, 0.773–0.931). In the subgroup analysis by sex, the AUROC was 0.926 (95% CI, 0.913–0.939) in females and 0.909 (95% CI, 0.897–0.921) in males.

**Table 4.**
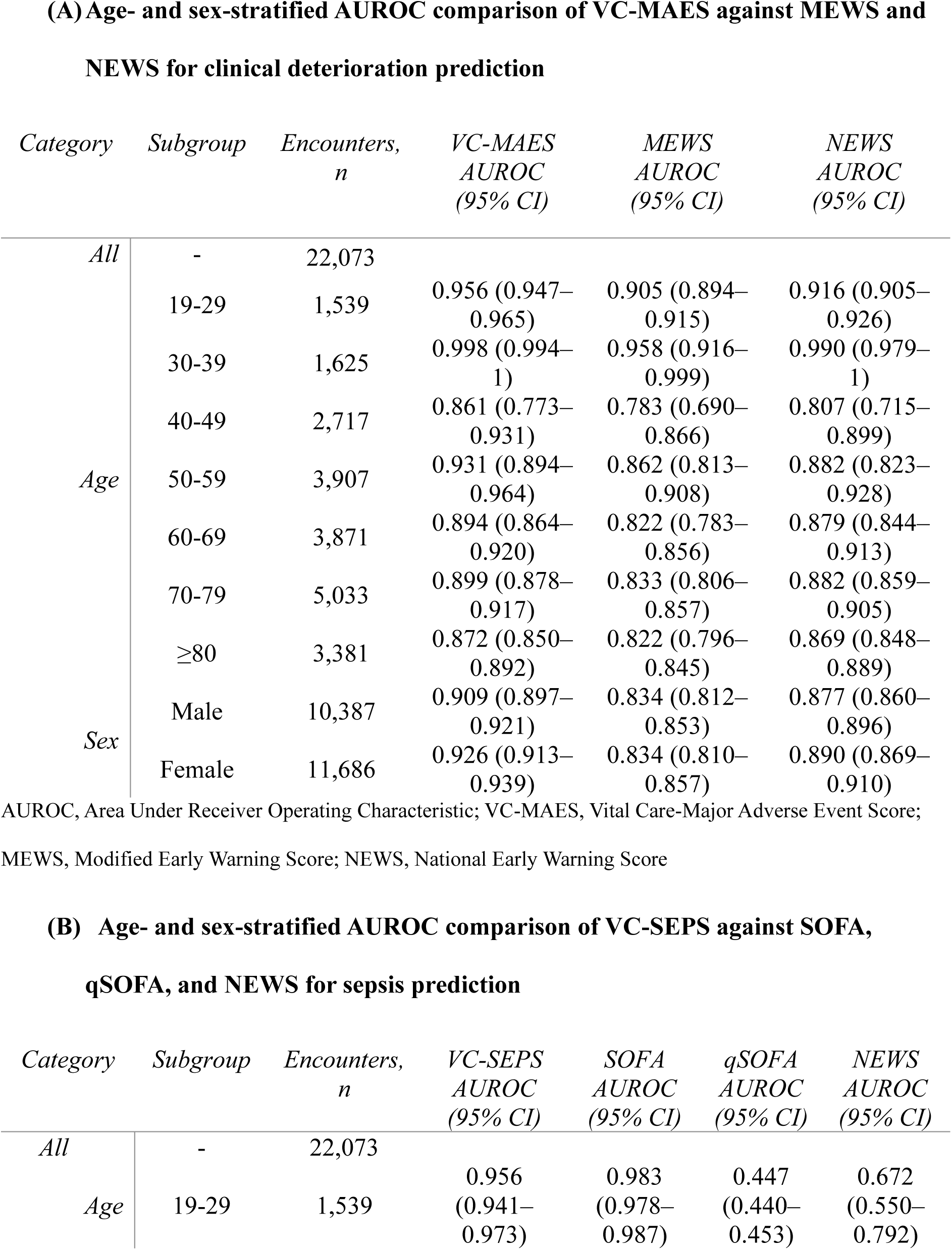
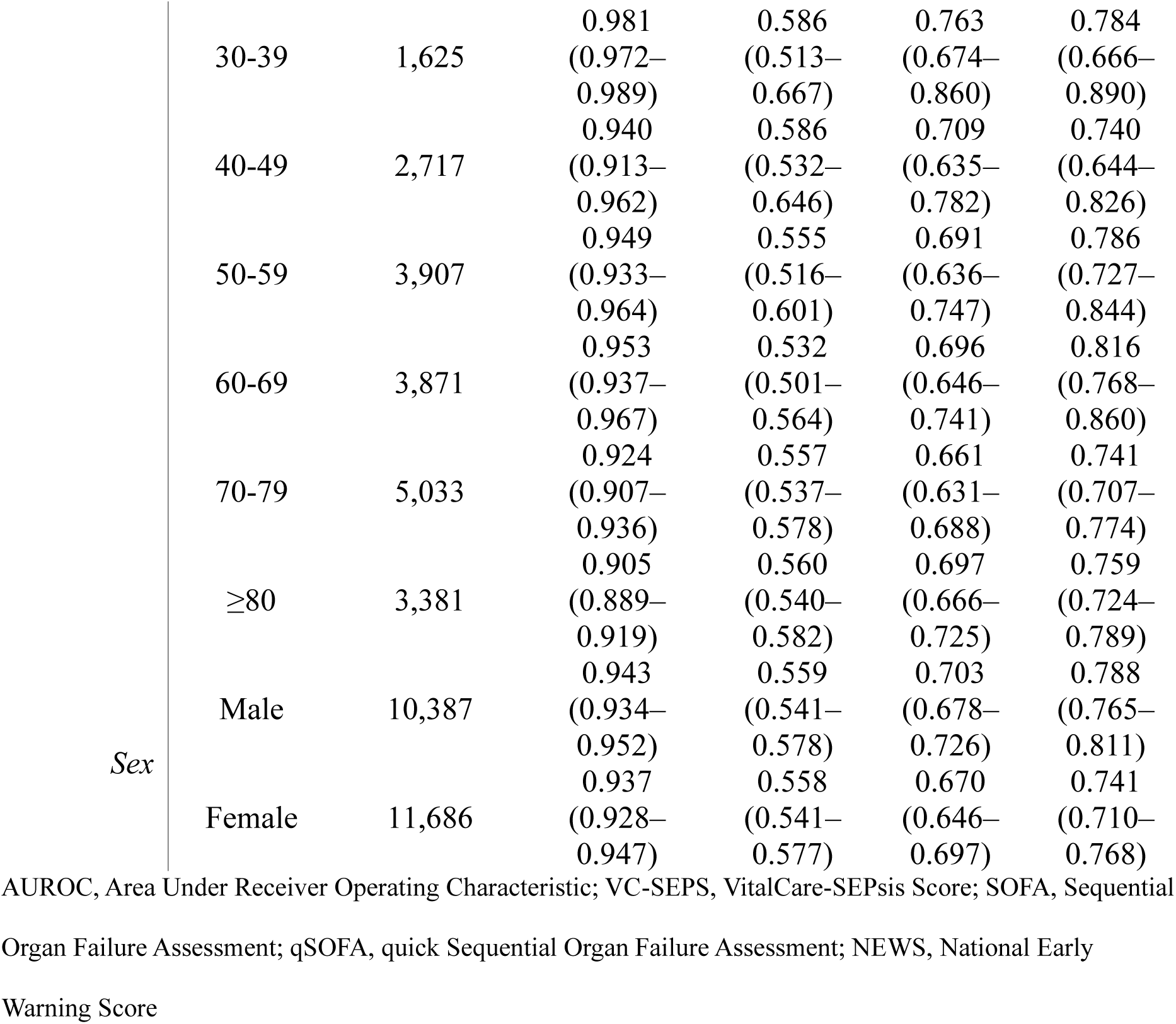
Subgroup Analyses of Model Performance by Age and Sex.

For sepsis prediction (Table 4B), the VC-SEPS achieved AUROCs of 0.905–0.981 across age groups, exceeding those of the SOFA (0.532–0.983), qSOFA (0.447–0.763), and NEWS (0.672–0.816) in all but one subgroup. In patients aged 20–29 years, the AUROC for VC-SEPS (0.956; 95% CI, 0.941–0.973) was slightly lower than that for SOFA (0.983; 95% CI, 0.978–0.987). By sex, the AUROC was 0.943 (95% CI, 0.934–0.952) in males and 0.937 (95% CI, 0.928–0.947) in females.

### Model Calibration

Calibration assessment in the external validation cohort is illustrated in **Figure 4**. The original VC-MAES demonstrated an expected calibration error (ECE) of 0.418, which improved markedly to 0.025 after calibration (Figure 4A). Similarly, the original VC-SEPS had an ECE of 0.306, decreasing to 0.006 post-calibration (Figure 4B). These results indicate substantial alignment between predicted and observed event probabilities following calibration.

**Fig 4.**
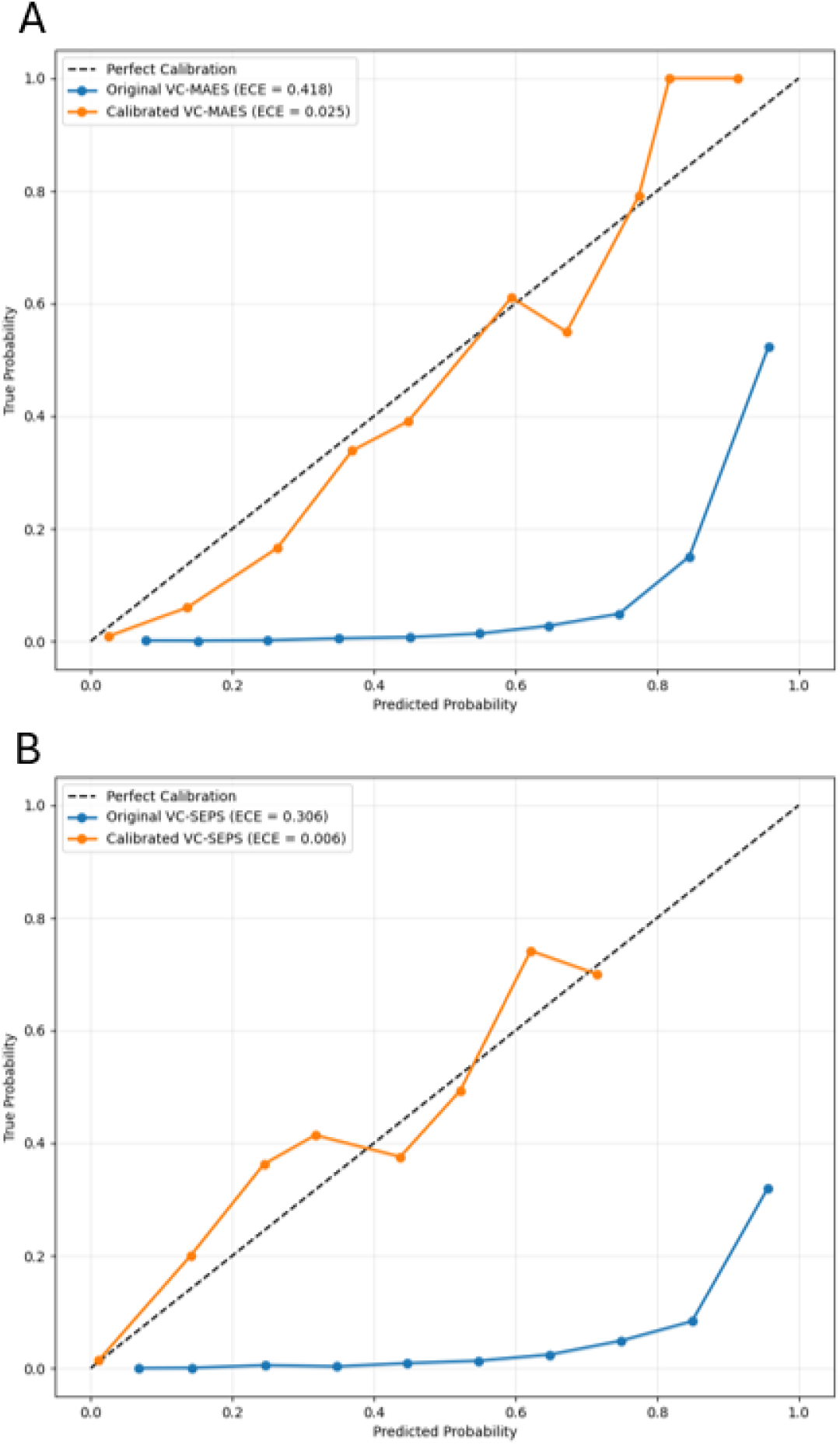
Calibration plot in the external validation cohort illustrating the observed and expected outcome rates across the VC-MAES and VC-SEPS score. VC-MAES (A) and VC-SEPS (B) calibration curve. The figure compares the actual probability (Y-axis) to the model’s predicted probability (X-axis) for the outcome of deterioration within 6 h (VC-MAES) and 4 h (VC-SEPS) in the external validation cohort.

## Discussion

In the present study, we developed and externally validated two DL-based early warning models—the VC**-**MAES for predicting clinical deterioration within 6 h and the VC**-**SEPS for predicting sepsis onset within 4 h—using collected EHR data in GW settings. External validation is essential for assessing generalizability. In our study, the external validation cohort differed notably from the derivation cohort in outcome prevalence, with lower rates of ICU transfer (1.4% vs. 4.7%) and higher rates of in-hospital mortality (2.9% vs. 1.1%) and sepsis (3.7% vs. 2.2%). Despite these differences, both models maintained high discriminative performance, with the VC**-**MAES achieving an AUROC of 0.918 and the VC**-** SEPS an AUROC of 0.941, significantly outperforming conventional risk scores, supporting the robustness, low risk of overfitting, and potential generalizability of the models across diverse clinical settings.

In subgroup analyses, both the VC-MAES and VC-SEPS maintained high discriminative ability (AUROC > 0.86) across all age and sex categories, supporting model fairness. For sepsis prediction in the 19–29-year age group, the SOFA marginally outperformed VC-SEPS (0.983 vs. 0.956); however, the performance of SOFA declined significantly in other age groups, with AUROC values educing to 0.5–0.6, whereas the VC-SEPS maintained AUROC > 0.9 across all age categories. Additionally, the sepsis prevalence in the 19-29-year age group was only 0.26%, which is markedly lower than 3.7% in the overall external validation cohort and likely contributed to this result. Overall, these findings indicate consistent performance across demographic subgroups and support the fairness of both VC-MAES and VC-SEPS, with no evidence of bias.

Traditional EWS such as the MEWS and NEWS remain widely used in clinical practice due to their simplicity and ease of interpretation (55). However, their predictive performance is often suboptimal. A meta-analysis including 21 studies with 107,008 infected patients outside the ICU found a moderate pooled AUROC of 0.70 for in-hospital mortality prediction, with lower performance observed in older patients using the NEWS (AUROC 0.63), and even poorer results with systemic inflammatory response syndrome criteria (AUROC 0.60) (56). Despite these limitations, their continued widespread use highlights the need for more accurate and timely predictive approaches in GW settings.

Recent advances in AI have enabled the development of models that capture complex, non-linear relationships in clinical data, leading to improved early detection of deterioration events (22). For instance, DeepAISE, a DL-based model for sepsis prediction, achieved AUROCs of 0.87–0.90 for sepsis onset (57), whereas InSight, an ML-based model for early sepsis detection, reported an AUROCs of 0.92 (58), both outperforming traditional rule-based scores. Building on these developments, the VC-MAES and VC-SEPS apply DL architectures capable of modeling temporal dependencies in patient data, which likely contributed to their ability to more promptly and accurately predict deterioration and sepsis compared to existing scores. By leveraging continuously updated EHR data and capturing subtle temporal trajectories, the VC-MAES and VC-SEPS may provide clinicians with actionable lead-time for intervention. These findings support their integration into real-world clinical workflows, with the goal of enabling earlier recognition and timely therapeutic action for high-risk patients.

### Study limitations

Although our models demonstrated strong retrospective performance and successful external validation, several limitations warrant discussion. First, rigorous prospective validation is necessary to establish real-world clinical utility. Prior experience with commercial models such as the Epic Sepsis Model, has shown that strong retrospective metrics do not always translate to clinical utility, often due to limited generalizability and poor calibration (59). Although external validation in a separate cohort represents an important step, continuous recalibration and additional prospective studies across diverse hospital settings and populations are required. Additionally, continuous model updates will be necessary to maintain predictive accuracy amid evolving clinical practices, demographic shifts, and changes in hospital management strategies (60). Furthermore, prospective clinical studies demonstrating improved patient outcomes, such as reduced ICU transfers or mortality, are essential to confirm that these models lead to meaningful improvements in patient care beyond statistical prediction alone. These continued validation efforts will ensure that the predictive models remain reliable and broadly applicable across healthcare contexts.

Second, the outcome definitions used in this study present some limitations. Regarding clinical deterioration, we defined unplanned ICU transfers as direct transfers from a general ward to the ICU. However, this definition may not have captured all ICU transfers resulting from clinical deterioration. For sepsis, our current use of the ASE criteria likely identifies a more severely ill cohort (40). Although this approach effectively aligns with the Sepsis-3 criteria for automated surveillance, further studies are needed to evaluate model performance under alternative sepsis definitions to broaden applicability.

Another important consideration is the practical integration of these models into real-time clinical workflows, which requires careful management of infrastructure demands, continuous performance monitoring, and clinician trust. One known barrier to trust and adoption is the “black box” nature of DL models, which rely on complex and opaque computations, unlike traditional scoring systems with transparent logic (61). To address this, we plan to incorporate explainable artificial intelligence methodologies into future model iterations of the VC-MAES and VC-SEPS. These methods aim to enhance transparency by identifying the most influential clinical features and clarifying the rationale behind predictions, thereby supporting clinician trust and facilitating successful clinical adoption (62, 63).

The strengths of the VC-MAES and VC-SEPS models are evident, including superior predictive accuracy, potential for earlier clinical intervention, comprehensive data utilization, and fairness demonstrated through subgroup validation by age and sex. However, their successful clinical integration depends on addressing key challenges, including interpretability, clinical acceptance, infrastructure readiness, and continuous monitoring. Therefore, successful implementation of AI-based EWSs will require careful balancing of predictive complexity with interpretability and actionable clinical guidance.

## Conclusion

In this study, we successfully developed and externally validated two DL-based predictive models, the VC-MAES and VC-SEPS, for the early identification of clinical deterioration and sepsis, respectively. Our findings demonstrate that these models offer improved predictive performance compared to traditional EWSs, providing accurate and timely alerts.

These systems can enhance patient safety by enabling earlier interventions, reducing preventable adverse events, and mitigating the clinical and financial burden associated with patient deterioration. Although these models show promise, their full realization in practice depends on addressing key challenges such as rigorous prospective validation in diverse clinical settings, ensuring model interpretability for clinician trust, and seamless integration into existing healthcare workflows. Through these implementation strategies, AI-driven solutions like the VC-MAES and VC-SEPS can lead to meaningful improvements in clinical outcomes and overall patient safety.

## Data Availability

All data produced in the present study are available upon reasonable request to the authors.

## References

1. Churpek MM, Carey KA, Snyder A, Winslow CJ, Gilbert E, Shah NS, et al. Multicenter Development and Prospective Validation of eCARTv5: A Gradient-Boosted Machine-Learning Early Warning Score. Crit Care Explor. 2025;7(4):e1232.

2. Giese A, Khanam R, Nghiem S, Staines A, Rosemann T, Boes S, et al. Assessing the excess costs of the in-hospital adverse events covered by the AHRQ’s Patient Safety Indicators in Switzerland. PLOS ONE. 2024;19(2):e0285285.

3. Curtis K, Sivabalan P, Bedford DS, Considine J, D’Amato A, Shepherd N, et al. Treatments costs associated with inpatient clinical deterioration. Resuscitation. 2021;166:49–54.

4. Byrd TFt, Phelan TA, Ingraham NE, Langworthy BW, Bhasin A, Kc A, et al. Beyond Unplanned ICU Transfers: Linking a Revised Definition of Deterioration to Patient Outcomes. Crit Care Med. 2024;52(9):e439–e49.

5. Singer M, Deutschman CS, Seymour CW, Shankar-Hari M, Annane D, Bauer M, et al. The Third International Consensus Definitions for Sepsis and Septic Shock (Sepsis-3). Jama. 2016;315(8):801–10.

6. Churpek MM, Ingebritsen R, Carey KA, Rao SA, Murnin E, Qyli T, et al. Causes, Diagnostic Testing, and Treatments Related to Clinical Deterioration Events Among High-Risk Ward Patients. Critical Care Explorations. 2024;6(10):e1161.

7. Holdsworth LM, Kling SMR, Smith M, Safaeinili N, Shieh L, Vilendrer S, et al. Predicting and Responding to Clinical Deterioration in Hospitalized Patients by Using Artificial Intelligence: Protocol for a Mixed Methods, Stepped Wedge Study. JMIR Res Protoc. 2021;10(7):e27532.

8. Evans L, Rhodes A, Alhazzani W, Antonelli M, Coopersmith CM, French C, et al. Surviving sepsis campaign: international guidelines for management of sepsis and septic shock 2021. Intensive Care Med. 2021;47(11):1181–247.

9. Rüddel H, Thomas-Rüddel DO, Reinhart K, Bach F, Gerlach H, Lindner M, et al. Adverse effects of delayed antimicrobial treatment and surgical source control in adults with sepsis: results of a planned secondary analysis of a cluster-randomized controlled trial. Crit Care. 2022;26(1):51.

10. Tang F, Yuan H, Li X, Qiao L. Effect of delayed antibiotic use on mortality outcomes in patients with sepsis or septic shock: A systematic review and meta-analysis. Int Immunopharmacol. 2024;129:111616.

11. Churpek MM, Wendlandt B, Zadravecz FJ, Adhikari R, Winslow C, Edelson DP. Association between intensive care unit transfer delay and hospital mortality: A multicenter investigation. J Hosp Med. 2016;11(11):757–62.

12. Yealy DM, Mohr NM, Shapiro NI, Venkatesh A, Jones AE, Self WH. Early Care of Adults With Suspected Sepsis in the Emergency Department and Out-of-Hospital Environment: A Consensus-Based Task Force Report. Ann Emerg Med. 2021;78(1):1–19.

13. Islam KR, Prithula J, Kumar J, Tan TL, Reaz MBI, Sumon MSI, et al. Machine Learning-Based Early Prediction of Sepsis Using Electronic Health Records: A Systematic Review. J Clin Med. 2023;12(17).

14. Kim S, Kim TH. The association between nurse staffing level and length of stay in general ward and intensive care unit in Korea. Applied nursing research: ANR. 2022;63:151558.

15. Rajendran G, Tjen C, Hutchinson S, Fletcher S. Are general wards sufficiently staffed to care for level 1 patients? Critical Care. 2013;17(Suppl 2):P506.

16. Churpek MM, Wendlandt B, Zadravecz FJ, Adhikari R, Winslow C, Edelson DP. Association Between ICU Transfer Delay and Hospital Mortality: A Multicenter Investigation. Journal of hospital medicine. 2016;11(11):757–62.

17. Barwise A, Thongprayoon C, Gajic O, Jensen J, Herasevich V, Pickering BW. Delayed Rapid Response Team Activation Is Associated With Increased Hospital Mortality, Morbidity, and Length of Stay in a Tertiary Care Institution. Crit Care Med. 2016;44(1):54–63.

18. Gupta S, Green C, Subramaniam A, Zhen LD, Low E, Tiruvoipati R. The impact of delayed rapid response call activation on patient outcomes. J Crit Care. 2017;41:86–90.

19. Liu V, Kipnis P, Rizk NW, Escobar GJ. Adverse outcomes associated with delayed intensive care unit transfers in an integrated healthcare system. J Hosp Med. 2012;7(3):224–30.

20. Maharaj R, Raffaele I, Wendon J. Rapid response systems: a systematic review and meta-analysis. Critical Care. 2015;19(1):254.

21. McGaughey J, Fergusson DA, Van Bogaert P, Rose L. Early warning systems and rapid response systems for the prevention of patient deterioration on acute adult hospital wards. The Cochrane Database of Systematic Reviews. 2021;2021(11):CD005529.

22. Gerry S, Bonnici T, Birks J, Kirtley S, Virdee PS, Watkinson PJ, et al. Early warning scores for detecting deterioration in adult hospital patients: systematic review and critical appraisal of methodology. Bmj. 2020;369:m1501.

23. Downey CL, Tahir W, Randell R, Brown JM, Jayne DG. Strengths and limitations of early warning scores: A systematic review and narrative synthesis. Int J Nurs Stud. 2017;76:106–19.

24. Moor M, Rieck B, Horn M, Jutzeler CR, Borgwardt K. Early Prediction of Sepsis in the ICU Using Machine Learning: A Systematic Review. Front Med (Lausanne). 2021;8:607952.

25. Muralitharan S, Nelson W, Di S, McGillion M, Devereaux PJ, Barr NG, et al. Machine Learning-Based Early Warning Systems for Clinical Deterioration: Systematic Scoping Review. J Med Internet Res. 2021;23(2):e25187.

26. Nwanosike EM, Conway BR, Merchant HA, Hasan SS. Potential applications and performance of machine learning techniques and algorithms in clinical practice: A systematic review. Int J Med Inform. 2022;159:104679.

27. Wang Z, Wang W, Sun C, Li J, Xie S, Xu J, et al. A methodological systematic review of validation and performance of sepsis real-time prediction models. NPJ Digit Med. 2025;8(1):190.

28. Reyna MA, Josef CS, Jeter R, Shashikumar SP, Westover MB, Nemati S, et al. Early Prediction of Sepsis From Clinical Data: The PhysioNet/Computing in Cardiology Challenge 2019. Crit Care Med. 2020;48(2):210–7.

29. Beneyto-Ripoll C, Palazón-Bru A, Llópez-Espinós P, Martínez-Díaz AM, Gil-Guillén VF, de Los Ángeles Carbonell-Torregrosa M. A critical appraisal of the prognostic predictive models for patients with sepsis: Which model can be applied in clinical practice? Int J Clin Pract. 2021;75(8):e14044.

30. Mittermaier M, Raza MM, Kvedar JC. Bias in AI-based models for medical applications: challenges and mitigation strategies. NPJ Digit Med. 2023;6(1):113.

31. de Hond AAH, Steyerberg EW, van Calster B. Interpreting area under the receiver operating characteristic curve. Lancet Digit Health. 2022;4(12):e853–e5.

32. Obuchowski NA, McClish DK. Sample size determination for diagnostic accuracy studies involving binormal ROC curve indices. Statistics in Medicine. 1997;16(13):1529–42.

33. Hajian-Tilaki K. Sample size estimation in diagnostic test studies of biomedical informatics. Journal of Biomedical Informatics. 2014;48:193–204.

34. Reese J, Deakyne SJ, Blanchard A, Bajaj L. Rate of Preventable Early Unplanned Intensive Care Unit Transfer for Direct Admissions and Emergency Department Admissions. Hospital Pediatrics. 2015;5(1):27–34.

35. Nolan JP, Berg RA, Andersen LW, Bhanji F, Chan PS, Donnino MW, et al. Cardiac Arrest and Cardiopulmonary Resuscitation Outcome Reports: Update of the Utstein Resuscitation Registry Template for In-Hospital Cardiac Arrest: A Consensus Report From a Task Force of the International Liaison Committee on Resuscitation (American Heart Association, European Resuscitation Council, Australian and New Zealand Council on Resuscitation, Heart and Stroke Foundation of Canada, InterAmerican Heart Foundation, Resuscitation Council of Southern Africa, Resuscitation Council of Asia). Circulation. 2019;140(18):e746–e57.

36. Churpek MM, Yuen TC, Edelson DP. Predicting clinical deterioration in the hospital: the impact of outcome selection. Resuscitation. 2013;84(5):564–8.

37. Haegdorens F, Van Bogaert P, Roelant E, De Meester K, Misselyn M, Wouters K, et al. The introduction of a rapid response system in acute hospitals: A pragmatic stepped wedge cluster randomised controlled trial. Resuscitation. 2018;129:127–34.

38. Rhee C, Dantes R, Epstein L, Murphy DJ, Seymour CW, Iwashyna TJ, et al. Incidence and Trends of Sepsis in US Hospitals Using Clinical vs Claims Data, 2009-2014. JAMA. 2017;318(13):1241–9.

39. Centers for Disease Control and Prevention. Hospital toolkit for adult sepsis surveillance Atlanta, GA: U.S. Department of Health and Human Services; 2018 [Available from: https://stacks.cdc.gov/view/cdc/132387/cdc_132387_DS1.pdf.

40. Rhee C, Zhang Z, Kadri SS, Murphy DJ, Martin GS, Overton E, et al. Sepsis Surveillance Using Adult Sepsis Events Simplified eSOFA Criteria Versus Sepsis-3 Sequential Organ Failure Assessment Criteria. Crit Care Med. 2019;47(3):307–14.

41. Shappell CN, Klompas M, Rhee C. Surveillance Strategies for Tracking Sepsis Incidence and Outcomes. J Infect Dis. 2020;222(Suppl 2):S74–s83.

42. Lee SY, Park MH, Oh DK, Lim CM. Validation of Adult Sepsis Event and Epidemiologic Analysis of Sepsis Prevalence and Mortality Using Adult Sepsis Event’s Electronic Health Records-Based Sequential Organ Failure Assessment Criteria: A Single-Center Study in South Korea. Crit Care Med. 2024;52(8):1173–82.

43. Rhee C, Kadri S, Huang SS, Murphy MV, Li L, Platt R, et al. Objective Sepsis Surveillance Using Electronic Clinical Data. Infection Control and Hospital Epidemiology. 2016;37(2):163–71.

44. Rhee C, Dantes R, Epstein L, Murphy DJ, Seymour CW, Iwashyna TJ, et al. Incidence and Trends of Sepsis in US Hospitals Using Clinical vs Claims Data, 2009-2014. JAMA. 2017;318(13):1241–9.

45. Rhee C, Dantes RB, Epstein L, Klompas M. Using objective clinical data to track progress on preventing and treating sepsis: CDC’s new ’Adult Sepsis Event’ surveillance strategy. BMJ quality & safety. 2019;28(4):305–9.

46. Lancia G, Varkila MRJ, Cremer OL, Spitoni C. Two-step interpretable modeling of ICU-AIs. Artificial Intelligence in Medicine. 2024;151:102862.

47. Gandin I, Scagnetto A, Romani S, Barbati G. Interpretability of time-series deep learning models: A study in cardiovascular patients admitted to Intensive care unit. Journal of Biomedical Informatics. 2021;121:103876.

48. Sun M, Engelhard MM, Bedoya AD, Goldstein BA. Incorporating informatively collected laboratory data from EHR in clinical prediction models. BMC Medical Informatics and Decision Making. 2024;24(1):206.

49. Steyerberg EW, Vickers AJ, Cook NR, Gerds T, Gonen M, Obuchowski N, et al. Assessing the performance of prediction models: a framework for traditional and novel measures. Epidemiology. 2010;21(1):128–38.

50. Ozenne B, Subtil F, Maucort-Boulch D. The precision--recall curve overcame the optimism of the receiver operating characteristic curve in rare diseases. J Clin Epidemiol. 2015;68(8):855–9.

51. Yuan Y, Su W, Zhu M. Threshold-free measures for assessing the performance of medical screening tests. Front Public Health. 2015;3:57.

52. Smith GB, Prytherch DR, Meredith P, Schmidt PE, Featherstone PI. The ability of the National Early Warning Score (NEWS) to discriminate patients at risk of early cardiac arrest, unanticipated intensive care unit admission, and death. Resuscitation. 2013;84(4):465–70.

53. Guan G, Lee CMY, Begg S, Crombie A, Mnatzaganian G. The use of early warning system scores in prehospital and emergency department settings to predict clinical deterioration: A systematic review and meta-analysis. PLoS One. 2022;17(3):e0265559.

54. Chua WL, Rusli KDB, Aitken LM. Early warning scores for sepsis identification and prediction of in-hospital mortality in adults with sepsis: A systematic review and meta-analysis. J Clin Nurs. 2024;33(6):2005–18.

55. Kramer AA, Sebat F, Lissauer M. A review of early warning systems for prompt detection of patients at risk for clinical decline. J Trauma Acute Care Surg. 2019;87(1S Suppl 1):S67–s73.

56. Zhang K, Zhang X, Ding W, Xuan N, Tian B, Huang T, et al. National Early Warning Score Does Not Accurately Predict Mortality for Patients With Infection Outside the Intensive Care Unit: A Systematic Review and Meta-Analysis. Front Med (Lausanne). 2021;8:704358.

57. Shashikumar SP, Josef CS, Sharma A, Nemati S. DeepAISE - An interpretable and recurrent neural survival model for early prediction of sepsis. Artif Intell Med. 2021;113:102036.

58. Mao Q, Jay M, Hoffman JL, Calvert J, Barton C, Shimabukuro D, et al. Multicentre validation of a sepsis prediction algorithm using only vital sign data in the emergency department, general ward and ICU. BMJ Open. 2018;8(1):e017833.

59. Wong A, Otles E, Donnelly JP, Krumm A, McCullough J, DeTroyer-Cooley O, et al. External Validation of a Widely Implemented Proprietary Sepsis Prediction Model in Hospitalized Patients. JAMA Intern Med. 2021;181(8):1065–70.

60. Davis SE, Greevy RA, Jr., Lasko TA, Walsh CG, Matheny ME. Detection of calibration drift in clinical prediction models to inform model updating. J Biomed Inform. 2020;112:103611.

61. Shortliffe EH, Sepúlveda MJ. Clinical Decision Support in the Era of Artificial Intelligence. Jama. 2018;320(21):2199–200.

62. Markus AF, Kors JA, Rijnbeek PR. The role of explainability in creating trustworthy artificial intelligence for health care: A comprehensive survey of the terminology, design choices, and evaluation strategies. J Biomed Inform. 2021;113:103655.

63. Jung J, Kang S, Choi J, El-Kareh R, Lee H, Kim H. Evaluating the impact of explainable AI on clinicians’ decision-making: A study on ICU length of stay prediction. Int J Med Inform. 2025;201:105943.

