## Supplemental Table 1 for "Development and Validation of VC-MAES and VC-SEPS: Deep Learning-Based Early Warning Systems for Hospitalized Patients"

**Table S1. Normal reference midpoints used for missing value imputation**

| Feature | Imputation normal value |
| --- | --- |
| SpO2 (%) | 100 |
| Glasgow Coma Scale | 15 |
| Total bilirubin (mg/dL) | 0.6 |
| Lactate (mmol/L) | 0.7 |
| Creatinine (mg/dL) | 0.8 |
| Platelets (10^9^/$\mathbf{L}$) | 300 |
| pH | 7.4 |
| Sodium (mmol/L) | 140 |
| Potassium (mmol/L) | 4.2 |
| Hematocrit (%) | 45 |
| White blood cell count (10^9^/$\mathbf{L}$) | 7 |
| HCO_3_^-^ | 24 |
| C-reactive protein (mg/dL) | 1.5 |
