## Supplemental Table 2 for "Development and Validation of VC-MAES and VC-SEPS: Deep Learning-Based Early Warning Systems for Hospitalized Patients"

|  | Yonsei Severance Hospital | | NHIS ILSAN Hospital | |
| --- | --- | --- | --- | --- |
| **Total N** | **357,025** | | **22,110** | |
| **Feature** | **Missing Count** | **Missing Rate** | **Missing Count** | **Missing Rate** |
| SBP | 0 | 0 % | 0 | 0 % |
| DBP | 0 | 0 % | 0 | 0 % |
| HR | 0 | 0 % | 0 | 0 % |
| RR | 0 | 0 % | 0 | 0 % |
| Body temperature | 0 | 0 % | 0 | 0 % |
| Oxygen saturation | 234,464 | 65.7 % | 10,048 | 45.4 % |
| Glasgow  Coma Scale | 44,952 | 12.6 % | 21,180 | 95.8 % |
| Total Bilirubin | 283,155 | 79.3 % | 7,485 | 33.9 % |
| Lactate | 351,729 | 98.5 % | 21,938 | 99.2 % |
| pH | 280,195 | 78.5 % | 22,046 | 99.7 % |
| Sodium | 95,211 | 26.7 % | 4,028 | 18.2 % |
| Potassium | 95,189 | 26.7 % | 4,029 | 18.2 % |
| Creatinine | 229,403 | 64.3 % | 4,505 | 20.4 % |
| Hematocrit | 75,235 | 21.1 % | 2,946 | 13.3 % |
| WBC | 75,235 | 21.1 % | 2,946 | 13.3 % |
| Bicarbonate | 280,195 | 78.5 % | 22,046 | 99.7 % |
| Platelet | 80,045 | 22.4 % | 2,946 | 13.3 % |
| CRP | 208,004 | 58.3 % | 22,084 | 99.9 % |

**Table S2. Missing rate**

SBP, Systolic Blood Pressure; DBP, Diastolic Blood Pressure; HR, Heart rate; RR, Respiratory Rate; CRP, C-reactive protein; WBC, White Blood Cell
