## Supplemental Table 3 for "Development and Validation of VC-MAES and VC-SEPS: Deep Learning-Based Early Warning Systems for Hospitalized Patients"

**Table S3. Performance at Key Cut-off Values in the External Validation Cohort**

(A) **VC-MAES: Prediction of clinical deterioration within 6 h**

| **Score** | **Cut-off value** | **Sensitivity** | **Specificity** | **NPV** | **PPV** | **F1-score** |
| --- | --- | --- | --- | --- | --- | --- |
| VC-MAES | 1 | 0.994 (0.989-0.998) | 0.217 (0.212-0.222) | 0.999 (0.998-1.000) | 0.048 (0.045-0.051) | 0.357 (0.350-0.363) |
|  | 5 | 0.890 (0.872-0.909) | 0.755 (0.750-0.761) | 0.994 (0.993-0.995) | 0.126 (0.118-0.134) | 0.817 (0.809-0.826) |
|  | 10 | 0.798 (0.774-0.820) | 0.882 (0.878-0.886) | 0.991 (0.990-0.992) | 0.210 (0.198-0.223) | 0.838 (0.824-0.850) |
|  | 15 | 0.734 (0.706-0.759) | 0.933 (0.930-0.936) | 0.989 (0.988-0.990) | 0.302 (0.286-0.320) | 0.822 (0.804-0.837) |
|  | 20 | 0.678 (0.649-0.707) | 0.956 (0.954-0.958) | 0.987 (0.986-0.988) | 0.378 (0.356-0.399) | 0.793 (0.773-0.813) |
|  | 25 | 0.646 (0.617-0.675) | 0.964 (0.962-0.966) | 0.986 (0.984-0.987) | 0.414 (0.390-0.439) | 0.773 (0.752-0.794) |
|  | 30 | 0.595 (0.564-0.624) | 0.973 (0.972-0.975) | 0.984 (0.982-0.985) | 0.468 (0.443-0.495) | 0.738 (0.715-0.761) |
|  | 35 | 0.542 (0.513-0.574) | 0.981 (0.979-0.982) | 0.982 (0.980-0.983) | 0.524 (0.495-0.553) | 0.698 (0.673-0.724) |
|  | 40 | 0.500 (0.471-0.529) | 0.984 (0.982-0.985) | 0.980 (0.979-0.982) | 0.546 (0.516-0.576) | 0.663 (0.637-0.688) |
|  | 45 | 0.468 (0.442-0.500) | 0.986 (0.985-0.988) | 0.979 (0.977-0.981) | 0.575 (0.543-0.609) | 0.635 (0.610-0.663) |
|  | 50 | 0.392 (0.362-0.421) | 0.990 (0.989-0.991) | 0.976 (0.975-0.978) | 0.614 (0.576-0.652) | 0.562 (0.530-0.590) |
|  | 55 | 0.388 (0.356-0.414) | 0.990 (0.989-0.992) | 0.976 (0.974-0.978) | 0.614 (0.576-0.650) | 0.558 (0.524-0.584) |
|  | 60 | 0.275 (0.249-0.302) | 0.993 (0.992-0.994) | 0.972 (0.970-0.974) | 0.615 (0.571-0.659) | 0.430 (0.398-0.463) |
|  | 65 | 0.221 (0.196-0.246) | 0.995 (0.994-0.996) | 0.970 (0.968-0.972) | 0.650 (0.597-0.702) | 0.362 (0.327-0.395) |
|  | 70 | 0.092 (0.074-0.109) | 0.999 (0.999-0.999) | 0.965 (0.963-0.967) | 0.808 (0.740-0.882) | 0.168 (0.137-0.197) |
|  | 75 | 0.088 (0.071-0.105) | 0.999 (0.999-1.000) | 0.965 (0.963-0.968) | 0.823 (0.755-0.896) | 0.161 (0.132-0.189) |
|  | 80 | 0.009 (0.004-0.015) | 1.000 (1.000-1.000) | 0.962 (0.960-0.965) | 1.000 (1.000-1.000) | 0.019 (0.008-0.030) |
|  | 85 | 0.002 (0.000-0.005) | 1.000 (1.000-1.000) | 0.962 (0.960-0.964) | 1.000 (0.000-1.000) | 0.004 (0.000-0.010) |
|  | 90 | 0.002 (0.000-0.005) | 1.000 (1.000-1.000) | 0.962 (0.960-0.964) | 1.000 (0.000-1.000) | 0.004 (0.000-0.009) |
|  | 95 | 0.000 (0.000-0.000) | 1.000 (1.000-1.000) | 0.962 (0.960-0.964) | 0.000 (0.000-0.000) | 0.000 (0.000-0.000) |
|  | 100 | 0.000 (0.000-0.000) | 1.000 (1.000-1.000) | 0.962 (0.960-0.964) | 0.000 (0.000-0.000) | 0.000 (0.000-0.000) |

VC-MAES, VitalCare-Major Adverse Event Score; NPV, Negative Predictive Value; PPV, Positive Predictive Value

(B) **VC-SEPS: Prediction of sepsis within 4 h**

| **Score** | **Cut-off value** | **Sensitivity** | **Specificity** | **NPV** | **PPV** | **F1-score** |
| --- | --- | --- | --- | --- | --- | --- |
| VC-SEPS | 1 | 0.951 (0.935-0.966) | 0.754 (0.749-0.759) | 0.998 (0.997-0.999) | 0.107 (0.099-0.114) | 0.841 (0.834-0.847) |
|  | 5 | 0.764 (0.734-0.794) | 0.923 (0.920-0.926) | 0.992 (0.991-0.993) | 0.234 (0.219-0.249) | 0.836 (0.818-0.854) |
|  | 10 | 0.571 (0.538-0.604) | 0.970 (0.967-0.972) | 0.987 (0.985-0.988) | 0.367 (0.339-0.393) | 0.719 (0.692-0.744) |
|  | 15 | 0.511 (0.474-0.544) | 0.976 (0.974-0.978) | 0.985 (0.983-0.986) | 0.400 (0.369-0.430) | 0.671 (0.638-0.699) |
|  | 20 | 0.474 (0.440-0.508) | 0.982 (0.980-0.983) | 0.984 (0.982-0.985) | 0.443 (0.410-0.473) | 0.639 (0.608-0.669) |
|  | 25 | 0.395 (0.363-0.428) | 0.986 (0.985-0.987) | 0.981 (0.980-0.983) | 0.466 (0.428-0.505) | 0.565 (0.531-0.597) |
|  | 30 | 0.352 (0.319-0.384) | 0.988 (0.987-0.990) | 0.980 (0.979-0.982) | 0.480 (0.438-0.519) | 0.519 (0.482-0.553) |
|  | 35 | 0.232 (0.204-0.261) | 0.993 (0.992-0.994) | 0.977 (0.975-0.978) | 0.521 (0.474-0.571) | 0.376 (0.338-0.414) |
|  | 40 | 0.226 (0.197-0.254) | 0.994 (0.993-0.995) | 0.977 (0.975-0.978) | 0.526 (0.477-0.575) | 0.368 (0.329-0.404) |
|  | 45 | 0.163 (0.137-0.189) | 0.997 (0.996-0.998) | 0.975 (0.973-0.977) | 0.628 (0.556-0.694) | 0.280 (0.240-0.318) |
|  | 50 | 0.156 (0.129-0.182) | 0.997 (0.997-0.998) | 0.975 (0.973-0.976) | 0.642 (0.580-0.714) | 0.269 (0.229-0.308) |
|  | 55 | 0.116 (0.096-0.139) | 0.999 (0.998-0.999) | 0.973 (0.972-0.975) | 0.715 (0.636-0.791) | 0.208 (0.176-0.244) |
|  | 60 | 0.112 (0.092-0.133) | 0.999 (0.998-0.999) | 0.973 (0.972-0.975) | 0.729 (0.651-0.806) | 0.201 (0.168-0.235) |
|  | 65 | 0.043 (0.030-0.056) | 0.999 (0.999-1.000) | 0.971 (0.969-0.973) | 0.720 (0.585-0.833) | 0.082 (0.057-0.106) |
|  | 70 | 0.033 (0.022-0.046) | 1.000 (0.999-1.000) | 0.971 (0.969-0.973) | 0.700 (0.553-0.830) | 0.064 (0.043-0.088) |
|  | 75 | 0.000 (0.000-0.000) | 1.000 (1.000-1.000) | 0.970 (0.968-0.972) | 0.000 (0.000-0.000) | 0.000 (0.000-0.000) |
|  | 80 | 0.000 (0.000-0.000) | 1.000 (1.000-1.000) | 0.970 (0.968-0.972) | 0.000 (0.000-0.000) | 0.000 (0.000-0.000) |
|  | 85 | 0.000 (0.000-0.000) | 1.000 (1.000-1.000) | 0.970 (0.968-0.972) | 0.000 (0.000-0.000) | 0.000 (0.000-0.000) |
|  | 90 | 0.000 (0.000-0.000) | 1.000 (1.000-1.000) | 0.970 (0.968-0.972) | 0.000 (0.000-0.000) | 0.000 (0.000-0.000) |
|  | 95 | 0.000 (0.000-0.000) | 1.000 (1.000-1.000) | 0.970 (0.968-0.972) | 0.000 (0.000-0.000) | 0.000 (0.000-0.000) |
|  | 100 | 0.000 (0.000-0.000) | 1.000 (1.000-1.000) | 0.970 (0.968-0.972) | 0.000 (0.000-0.000) | 0.000 (0.000-0.000) |

VC-SEPS, VitalCare-Sepsis Score; NPV, Negative Predictive Value; PPV, Positive Predictive Value
